# Rates of osmotic demyelination after rapid correction of sodium in hyponatremia, a multicenter cohort study of patients hospitalized with hyponatremia

**DOI:** 10.1101/2022.08.29.22278419

**Authors:** Thomas E MacMillan, Saeha Shin, Joel Topf, Janice Kwan, Adina Weinerman, Terence Tang, Afsaneh Raissi, Radha Koppula, Fahad Razak, Amol A Verma, Michael Fralick

## Abstract

**Background:** Osmotic demyelination syndrome (ODS) is a rare but devastating complication of rapid correction of hyponatremia. Current guidelines recommend limiting the sodium correction rate to no more than 8 mmol/L per 24 hours, but this is based on expert opinion and small observational studies.

**Methods:** We conducted a multicenter cohort study of patients admitted into hospital with hyponatremia at five academic hospitals in Toronto between April 1, 2010 and December 31, 2019. We identified all adult patients with hyponatremia (sodium <130 mmol/L) based on their initial serum sodium measured on presentation to the emergency department. The primary outcome was the rate of ODS. ODS cases were identified using medical record review and neuroimaging results. The secondary outcome was the rate of rapid correction of sodium (>8 mmol/L in any 24-hour period).

**Results:** Our cohort included 21182 hospitalizations with hyponatremia. Approximately 50% were women, the average age was 68 years, and mean initial sodium was 124.6 mmol/L (SD 4.6) including 13.1% with sodium <120 mmol/L. Overall, rapid correction of sodium occurred in 3438 (17.9%) admissions. Despite the fact that 3438 experienced rapid correction, there were only 12 cases of ODS among our 21182 hospitalizations with hyponatremia. Cases of ODS had a markedly lower initial serum sodium (110.7 vs. 124.6 mmol/L), were younger (50 years vs 68 years), were more likely to have alcohol use disorder, and were more likely to have hypokalemia (58% vs 14%) compared to those without ODS.

**Conclusions:** In the large multicenter study of patients with hyponatremia, “rapid” overcorrection was common (N=3438, 17.9%) but ODS was rare (N=12, 0.06%). The initial serum sodium was markedly lower for those with ODS compared to those without. Taken together, these results suggest that the severity of hyponatremia is a more important risk factor for ODS then the rate of correction.

## INTRODUCTION

Rapid correction of hyponatremia is a risk factor for osmotic demyelination syndrome (ODS) which can lead to temporary or permanent neurologic injury.^1^ Although ODS is rare, current guidelines recommend limiting the sodium correction rate to no more than 8 mmol/L per 24 hours in all patients at high risk of ODS.^2^ This recommendation is based on expert opinion and observational studies. Two of the largest studies conducted to date each included approximately 1500 patients with hyponatremia and rates of ODS ranged from 0.28-0.53%; rapid correction occurred in 67-88% of ODS cases in these studies.^3,4^

More recently, some have advocated for applying the stringent sodium correction limit of <8 mmol/L to all patients with severe hyponatremia (sodium <120 mmol/L), regardless of individual risk factors for ODS.^5^ This recommendation is based on expert opinion and observational studies. There are potential drawbacks to applying more stringent sodium correction limits, such as more frequent bloodwork monitoring, prolonging the time to correction of hyponatremia, and increasing the length of stay in hospital.^3,6^ ODS can still occur even when the sodium correction rate is within the guideline-recommended range.^7^

While rapid correction is a risk factor for ODS, the relative importance of rapid correction relative to other risk factors for ODS such as initial sodium level is not clear. In this multicenter study, we sought to characterize the rate of ODS in patients hospitalized with hyponatremia based on initial sodium level. A second objective was to compare the rates of sodium correction between patients who did and did not develop ODS.

## METHODS

### Study setting

We conducted a multicenter cohort study of patients admitted into hospital with hyponatremia at five downtown academic hospitals in Toronto between April 1, 2010 and December 31, 2019. Toronto (population of 2.8 million) is the largest city in Canada and is among the most multicultural cities in the world. Hospital services are publicly insured for residents of Ontario with no out-of-pocket expenses incurred during a hospitalization. Research ethics board approval was obtained from all participating hospitals (Clinical Trials Ontario Study ID 1394, https://www.ctontario.ca/).

### Data Source

Data were collected from the GEMINI database, a hospital research collaborative.^8^ This database linked electronic patient data with administrative hospital data for all patients hospitalized under general internal medicine (GIM). Patients admitted to GIM generally present via the emergency department and elective/planned admissions are rare (<0.1% of all admissions). Patients were also included in the database if they presented to the emergency department and then went to the intensive care unit and were subsequently transferred to GIM. We included data from hospitals where laboratory and imaging results were readily available electronically. Additional data were manually extracted from the electronic admission note, medical imaging systems, and the electronic discharge summary.

### Study Population

We identified all admissions of adult patients with hyponatremia (sodium <130 mmol/L) based on their first serum sodium (“initial sodium”) measured on presentation to the emergency department. We included all patients who were subsequently admitted or transferred to GIM. We did not include patients who were discharged home from the emergency department without being admitted. Patients with hyponatremia are typically admitted to either GIM or critical care at the study hospitals. Subsequent admissions for the same patient during the study period were included if they met the inclusion criteria.

We chose to identify our cohort using laboratory data rather than diagnostic codes, because diagnostic codes have low sensitivity.^9^ We excluded patients with an initial blood glucose ≥25 mmol/L because hyperglycemia can result in low serum sodium values, and patients with a known history of diabetes insipidus because they may be taking desmopressin which can affect sodium correction rates (Appendix). We obtained data on comorbidities using Clinical Classifications Software Refined codes (Appendix).

### Study Outcomes

The primary outcome was the rate of osmotic demyelination syndrome (ODS) on the index admission. Possible ODS cases were identified using the results of neuroimaging (primarily MRI or CT) performed on the index admission. Radiologist reports for all brain MRI and CT scans performed after the initial sodium were searched using keywords to identify patients who may have had ODS (Appendix). In each case with positive keyword search, a trained reviewer reviewed the radiology report and identified a subset of cases for manual chart review. During the chart review, we examined the patient record for that admission and categorized the case as follows: no ODS, possible ODS, or definite ODS. This approach has been validated previously.^3,4^ Cases categorized as either possible or definite ODS were considered ODS cases for the main analysis and a subsequent sensitivity analysis included only definite ODS. Because reviewing imaging results alone may fail to identify patients with ODS, we also performed manual medical record review of all cases which had an ICD-10 discharge diagnosis of ODS (ICD-10 G37.2) even in the absence of supportive imaging. For any readmissions within 7 days of the index admission with hyponatremia, we searched the most responsible diagnosis for an ODS code to identify cases of ODS that may have developed after the hospitalization.

The secondary outcome was the rate of rapid correction of sodium. We report the percentage of patients who experienced rapid correction and the rates of ODS in this group. Prior studies have used sodium rapid correction as a surrogate outcome, since ODS is a rare outcome and difficult to diagnose.^10^ However, there is a lack of agreement about the optimal way to define rapid correction.^10^ We adapted a previously published method based on consensus guidelines and defined rapid correction as change in sodium >8 mmol/L in any 24-hour period from the initial sodium until either time of death, time of hospital discharge, or time of first sodium ≥130 mmol/L.^3^ We also calculated the rates of correction at 24 and 48 hours from the initial sodium by choosing values closest to that time point, up to 6 hours before or after the 24- and 48-hour time points respectively. We obtained urine sodium and osmolality results within 24 hours of the initial sodium.

### Statistical Analysis

Descriptive statistics were used to characterize patient-level characteristics and total length of hospitalization. To maintain confidentiality, results with 5 or fewer cases are reported as “≤5” as per the recommended practices for use of administrative health data in our province.^11^ We calculated the unadjusted overall cumulative incidence rate of ODS and the stratified cumulative incidence by initial sodium (<110 mmol/L, 110-119 mmol/L, ≥120 mmol/L). All analyses were performed using R (The R Foundation for Statistical Computing, 2021).

## Results

### Study Cohort

Between April 1, 2010, and December 31, 2019, there were 21610 admissions with an initial serum sodium of <130 mmol/L. We excluded 27 with a diagnosis of diabetes insipidus and 401 with an initial glucose ≥25 mmol/L. The remaining 21182 admissions represented 16034 unique patients, of which 2283 (10.8%) were admitted initially to critical care. Among all admissions, approximately 50% were women, the average age was 68 years, 2957 (14%) had initial potassium <3.5 mmol/L, and mean initial sodium was 124.6 mmol/L (SD 4.6) including 1.2% with an initial sodium <110 mmol/L, 11.9% from 110-119 mmol/L, and 86.9% ≥120 mmol/L. Overall, rapid correction (i.e., a change in serum sodium of >8 mmol/L over a 24-hour period) of sodium occurred in 3438 (17.9%) admissions.

#### Osmotic Demyelination Syndrome

A CT and MRI of the brain was performed in 29.6% and 5.9% of admissions respectively. Of the 1245 brain MRIs performed, 125 (10%) had a positive keyword search; manual review of the imaging report led to 33 of these (26%) undergoing further chart review. Of the 6273 brain CTs performed, 141 (2.2%) had a positive keyword search; manual review of the imaging report led to 10 of these (7%) undergoing further chart review.

Overall, there were 12 cases of ODS identified based on the chart review (0.06% of the cohort). Of these, 9 (75%) were categorized as definite ODS. Chart review of all admissions with an ICD-10 diagnosis code for ODS identified no new cases beyond those already included. Amongst the 998 patients readmitted within 7 days of hospital discharge, none had ODS listed as the most responsible diagnosis for the readmission, although a detailed clinical review of readmissions was not performed.

We observed _≤_5 cases of ODS in patients with an initial sodium of 125 mmol/L or greater. None of these cases experienced rapid correction (the change in sodium at 24 hours ranged from 2-8 mmol/L). However, in all these cases there was a prolonged period of hypernatremia after correction of the hyponatremia there were multiple known risk factors for ODS. In all cases, the sodium was >150 mmol/L for more than 24 hours, and the hypernatremia developed between 4-10 days post-admission.

#### Baseline characteristics by ODS status

Baseline characteristics by ODS status are shown in Table 1. Patients with ODS were younger (mean age 50 years, standard deviation (SD) 13 vs. no ODS 68 years SD 17), had a higher comorbidity burden (mean Elixhauser score 14.5, SD 5.9 vs. no ODS 9.0, SD 7.5), and a higher proportion of alcohol use history (between 8-42% versus no ODS 4.5%). ODS cases had lower initial sodium (mean 111 mmol/L, SD 10 vs. no ODS mean 125 mmol/L, SD 5), and in all ODS cases where urine sodium was measured it was <30 mmol/L. None of these had a diagnostic code for CHF or liver disease, meaning that the urine sodium of <30 mmol/L likely indicates hypovolemia. Overall, 9/4490 (0.2%) patients with urine sodium <30 mmol/L developed ODS. An initial sodium of <110 mmol/L was the most common presentation for patients developing ODS (7/12, 58.3%).

**Table 1.** Baseline characteristics by ODS status

|  | ODS | Not ODS |
| --- | --- | --- |
| n | 12 | 21170 |
| Age, mean (SD) | 50.25 (12.55) | 67.98 (16.76) |
| Male sex (%) | 6 (50.0) | 10550 (49.8) |
| Elixhauser comorbidity, mean (SD) | 14.50 (5.87) | 9.00 (7.51) |
| Pre-Admission Comorbidities* (%) |  |  |
| Cancer | 0 (0.0) | 1286 (6.1) |
| Alcohol use disorder | ≤5 (0-41.7) | 943 (4.5) |
| Diabetes | ≤5 (0-41.7) | 6120 (28.9) |
| Hypertension | ≤5 (0-41.7) | 8294 (39.2) |
| Heart failure | 0 (0.0) | 1804 (8.5) |
| Liver disease | 0 (0.0) | 1411 (6.7) |
| Renal Failure | 0 (0.0) | 2229 (10.5) |
| **Urine results on day 0** |  |  |
| Urine sodium (mmol/L) |  |  |
| ≤30 | 10 (83.3%) | 4658 (22.0) |
| >30 | ≤5 (0-41.7) | 5001 (23.6) |
| NA | ≤5 (0-41.7) | 11511 (54.4) |
| Urine osmolality (mOsm/kg) |  |  |
| mean (SD) | 264.7 (184.0) | 364.7 (172.7) |
| median [IQR] | 240.0 [142.3, 282.5] | 341.0 [238.0, 470.0] |
| NA | ≤5 (0-41.7) | 12972 (61.3) |
| **Serum results on day 0** |  |  |
| Sodium (mean (SD)) | 110.7 (10.4) | 124.6 (4.6) |
| Sodium (median [IQR]) | 110.0 [105.8, 114.8] | 126.0 [122.0, 128.0] |
| Sodium (mmol/L) |  |  |
| <110 | 7 (58.3) | 245 (1.2) |
| 110-119 | ≤5 (0-41.7) | 2528 (11.9) |
| ≥120 | ≤5 (0-41.7) | 18397 (86.9) |
| Serum osmolality (mOsm/kg) |  |  |
| mean (SD) | 235 (37.3) | 274.5 (27.1) |
| median [IQR] | 230.5 [207.8, 248.0] | 271.0 [258.0, 287.0] |
| NA | ≤5 (0-41.7) | 15837 (74.8) |
| Potassium (mmol/L) |  |  |
| ≤3.4 | 7 (58.3) | 2950 (13.9) |
| >3.4 | ≤5 (0-41.7) | 18202 (86.0) |
| NA | ≤5 (0-41.7) | 18 (0.1) |
| Alcohol level |  |  |
| Negative (<5 mmol/L) | 6 (50.0) | 2050 (9.7) |
| Positive (≥5 mmol/L) | ≤5 (0-41.7) | 378 (1.8) |
| NA | ≤5 (0-41.7) | 18742 (88.5) |

Legend - \*defined using CCSR codes, NA = not available

#### In-hospital management by ODS status

Patients with ODS were more likely to be admitted to critical care or receive DDAVP in hospital (Table 2). Each of the 5 hospital sites had at least one case of ODS, with _≤_5 cases per site. The low number of cases precludes comparing ODS rates by site.

**Table 2.** In-hospital Course and Outcomes by ODS status

|  | **ODS** | **Not ODS** |
| --- | --- | --- |
| **n** | **12** | **21170** |
| **Discharge to continuing care (%)** | 8 (66.7) | 3696 (17.5) |
| **Length of stay in acute care (days):** |  |  |
| mean (SD) | 40.3 (35.4) | 8.3 (12.1) |
| median [IQR] | 37.2 [14.2, 49.3] | 5.3 [2.8, 9.5] |
| **Died in hospital (%)** | ≤5 (0-41.7) | 1839 (8.7) |
| **Readmitted within 7 days (%)** | 0 (0.0) | 998 (4.7) |
| **Max. rate of sodium correction in 24h (mmol/L)** |  |  |
| ≤8 | 7 (58.3) | 15812 (74.7) |
| >8-<12 | ≤5 (0-41.7) | 2338 (11.0) |
| ≥12 | ≤5 (0-41.7) | 1095 (5.2) |
| NA | 0 (0.0) | 1925 (9.1) |
| **Change in serum sodium** |  |  |
| Sodium at 24 hours, mean (SD) - mmol/L | 7.2 (5.0) | 5.3 (4.1) |
| Sodium at 24 hours, median [IQR] - mmol/L | 7.5 [3.5, 9.0] | 5.0 [2.0, 8.0] |
| Serum Sodium result available at 24hr (%)* | 12 (100.0) | 11960 (56.5) |
| Sodium at 48 hours, mean (SD) - mmol/L | 11.3 (6.4) | 6.5 (4.8) |
| Sodium at 48 hours, median [IQR], mmol/L | 12.50 [8.3, 14.3] | 6.0 [3.0, 9.0] |
| Serum sodium result available at 48h (%) | 12 (100.0) | 9458 (44.7) |
| **Treatments** |  |  |
| DDAVP use in first 96 hours (%) | 6 (50.0) | 1278 (6.0) |
| **Imaging** |  |  |
| CT brain performed (%) | 11 (91.7) | 6262 (29.6) |
| Num. of days to CT, median [IQR] | 0.1 [0.1, 7.6] | 0.2 [0.1, 0.8] |
| MRI brain performed (%) | 11 (91.7) | 1234 (5.8) |
| Num. of days to MRI, median [IQR] | 21.1 [13.2, 34.9] | 3.4 [1.7, 7.0] |

\*This means that overall, only 56% of patients had a repeat sodium available at 24 plus or minus 6 hours

**Table 3.** Outcomes by Maximum Sodium Correction in 24 hours

| Maximum Sodium Correction<br>in 24 hours | Overall | ≤8 mmol/L | >8-<10<br>mmol/L | ≥10-<12<br>mmol/L | ≥12<br>mmol/L |
| --- | --- | --- | --- | --- | --- |
| n | 19257 | 15819 | 1036 | 1305 | 1097 |
| Age, mean (SD) | 67.97<br>(16.72) | 68.73<br>(16.52) | 65.87<br>(17.27) | 64.94<br>(16.90) | 62.56<br>(17.24) |
| Elixhauser comorbidity, mean<br>(SD) | 9.08<br>(7.49) | 9.34<br>(7.63) | 7.98<br>(6.67) | 8.05 (6.70) | 7.58<br>(6.76) |
| Sodium on day 0, mean (SD),<br>mmol/L | 124.35<br>(4.73) | 125.22<br>(3.94) | 121.57<br>(5.54) | 120.61<br>(5.72) | 118.85<br>(5.99) |
| Sodium on day 0 (mmol/L) |  |  |  |  |  |
| <110 | 252 (1.3) | 77 (0.5) | 39 (3.8) | 59 (4.5) | 77 (7.0) |
| 110-119 | 2511<br>(13.0) | 1335<br>(8.4) | 263<br>(25.4) | 414 (31.7) | 499<br>(45.5) |
| ≥120 | 16494<br>(85.7) | 14407<br>(91.1) | 734<br>(70.8) | 832 (63.8) | 521<br>(47.5) |
| Admission to ICU in first 72<br>hours (%) | 2244<br>(11.7) | 1535 (9.7) | 172 (16.6) | 267 (20.5) | 270<br>(24.6) |
| ≥1 DDAVP dose in first 96<br>hours (%) | 1280 (6.6) | 483 (3.1) | 175 (16.9) | 297 (22.8) | 325<br>(29.6) |
| Length of stay in acute care<br>(days): |  |  |  |  |  |
| mean (SD) | 8.4<br>(12.25) | 8.6<br>(11.43) | 8.0<br>(12.58) | 8.2 (19.99) | 7.6<br>(11.18) |
| median [IQR] | 5.4 [2.8,<br>9.7] | 5.5 [2.8,<br>9.9] | 4.8 [2.7,<br>9.0] | 4.9 [2.7,<br>8.8] | 4.8 [2.7,<br>8.5] |
| Readmitted within 7 days (%) | 903 (4.7) | 761 (4.8) | 43 (4.2) | 59 (4.5) | 40 (3.6) |
| Died in hospital (%) | 1630 (8.5) | 1459 (9.2) | 55 (5.3) | 63 (4.8) | 53 (4.8) |
| Discharge to continuing care<br>(%) | 3363<br>(17.5) | 2868<br>(18.1) | 151<br>(14.6) | 187 (14.3) | 157<br>(14.3) |
| ODS (%) | 12 (0.1) | 7 (0.0) | ≤5 (0-0.4) | ≤5 (0-0.4) | ≤5 (0-0.4) |

#### Outcomes by ODS status

Patients with ODS had longer length-of-stay and a higher proportion died in hospital or were discharged to continuing care. None of the patients in the ODS group was readmitted within 7 days.

#### Outcomes by rapid correction status

Among 19258 admissions with available data, 3436 (17.8%) had rapid correction of sodium (i.e., change of sodium >8 mmol/L in any 24-hour period). Overall rates of rapid correction ranged from 14-23% by individual hospital site. We could not assess for rapid correction in 1925 (9%) of admissions due to an inadequate number of sodium measurements or all of sodium measurements being too far apart in time (i.e., >24 hours apart).

Patients with rapid correction were more likely to be admitted initially to critical care (20.6% vs. 9.7% and receive DDAVP (23% vs 3%) but had similar readmission rates to those with no rapid correction (4.1 vs. 4.8%). Patients with an initial sodium <110 mmol/L underwent rapid correction 69.4% of the time (175/252), compared with 46.8% with sodium 110-119 mmol/L (1176/2155) and only 12.7% with sodium ≥120 mmol/L (1087/16494).

## DISCUSSION

In this large, multicenter study of over 20000 admissions of hospitalized patients with hyponatremia, there were 5 main findings. First, ODS was rare, occurring in 0.06% of admissions with sodium <130 mmol/L, regardless of the rate of sodium correction.

Second, low urine sodium (<30 mmol/L) and initial sodium <110 mmol/L were common in patients that developed ODS and may be important risk factors. Third, ODS developed in a small number of cases with initial sodium >125 mmol/L who developed subsequent hypernatremia.

Patients who developed ODS in our study had lower initial sodium levels and higher rates of hypokalemia and alcohol use, which are known risk factors for ODS.^2^ Patients with ODS had higher rates of DDAVP use in hospital, likely reflecting a reactive or rescue DDAVP strategy (i.e., administering DDAVP for impending sodium rapid correction, or after rapid correction has already occurred).^6^ The rates of ODS in this study were lower than the 0.25-0.5% incidence rate reported in recent studies,^3,4^ although those studies included patients with lower initial sodium levels (<120 mmol/L) who were likely at higher risk for ODS.

All the ODS patients with urine tests performed had urine sodium <30 mmol/L (with no history of CHF or liver disease) which likely reflects hypovolemia. However, only 0.2% of all patients with urine sodium <30 mmol/L developed ODS. It is unclear if urine sodium <30 mmol/L is an independent risk factor for ODS, since it may signify hypovolemia which can predispose to rapid sodium correction.

Fewer than 6 of the ODS cases in our study had an initial sodium level of >125 mmol/L, however it did occur. All these cases were complicated by a prolonged period of hypernatremia after correction of initial hyponatremia and had established risk factors for ODS; none experienced rapid correction. Our findings confirm the literature that ODS can develop in patients with mild hyponatremia or normonatremia, especially if other risk factors are present.^1^ A novel finding in our study is that hypernatremia occurring after correction of hyponatremia may be an important risk factor for ODS.

A significant proportion of our cohort (17.9%) had rapid correction of sodium (>8 mmol/L in 24 hours). Rapid correction was more frequent among patients with initial sodium <110 mmol/L, occurring in 69%. Despite the relatively high rate of rapid correction, ODS remained rare in our cohort, suggesting that in most cases, rapid correction of sodium alone does not lead to ODS, which is consistent with previous studies.^3,4^ Mortality and readmission rates were not elevated among patients that had rapid correction which suggests this group did not have an excess of undiagnosed ODS.

Importantly, among patients that developed ODS in this study, rapid correction occurred less than half, suggesting that other factors must be implicated in the development of ODS. In previous studies, rapid correction occurred at higher frequency with ODS (75-87%).^3,4^ The reason for the lower frequency of rapid correction in the ODS patients in this study is unclear, and the small number of cases in these studies makes it difficult to draw firm conclusions.

### Limitations

While a strength of this study is the inclusion of multiple sites, the 5 sites are all academic hospitals in a single city in Ontario, Canada, which may limit generalizability. Since the diagnoses contained in the GEMINI database are based on hospital administrative coding, we lacked certain predictors of ODS such as malnutrition, and the diagnostic codes that we used may have varying levels of accuracy. The study design precludes a definitive determination of the etiology of hyponatremia, especially given that a large proportion did not have urine electrolytes measured. Our database lacks information about intravenous fluid administration which can affect rates of sodium correction. The small number of ODS cases in the study precludes our ability to identify specific risk factors for ODS or define safe sodium correction targets. Finally, by using neuroimaging reports to identify cases of ODS, it is possible that we missed cases where neuroimaging was not performed. However, in an earlier study involving two of the study sites, a manual chart review of all patients who died did not identify any additional cases of ODS.^3^ It is possible we missed cases of ODS that presented on a subsequent admission, as we did not examine the readmissions in our study; however, none of the readmissions had a diagnostic code for ODS.

## CONCLUSION

In this multicenter study of 21182 hospital admissions for hyponatremia, we found that while rapid correction of sodium was common (17%), ODS was rare, occurring in only 0.06% of admissions. Patients with ODS had lower initial sodium and had a higher frequency of rapid sodium correction. An initial urine sodium of <30 mmol/L may also indicate increased risk of ODS in a patient with other risk factors. An initial sodium of <110 mmol/L was associated with high rates of rapid sodium correction. Our findings suggest that efforts to control the rate of sodium correction should be directed towards the highest risk patients, and that in the context of current clinical practice, patient risk factors appear to be at least as important as the rate of sodium correction for ODS.

## Data Availability

All data produced in the present study are available upon reasonable request to the authors.

## ACKNOWLEDGEMENTS

Hae Young Jung – coding

Yishan Guo – coding

Daniel Tamming – variable definition

## Appendix

**1) Inclusion and Exclusion criteria:**

|  | Overall |
| --- | --- |
| Inclusion step 1 : (admitted to GIM OR Critical Care through the emergency department (ED)) AND (discharged during study period (April 1, 2010 to December 31, 2019) with initial serum sodium (sNa) < 130 mmol/L) | 21610 |
| Exclusion step 1 : admitted initially to service that is not GIM or critical care | 21610 (0, 0%) |
| Exclusion step 2 : age < 18 | 21610 (0, 0%) |
| Exclusion step 3 : diagnosis of diabetes insipidus | 21583 (27, 0%) |
| Exclusion step 4 : severe hyperglycemia | 21182 (401, 2%) |

**2) Neuroimaging report text search keywords:**

- osmo*
- myelin*
- ods*
- cpm*
- pontine*

* = wildcard. All search terms were combined using OR

**3) Diagnostic codes**

| Diagnosis | CCSR CODES | Where to look for the codes |
| --- | --- | --- |
| Cancer | Cancer [11-47] (2018 version) | Current admission: pre-admit |
| Diabetes | END002, END003, END004, END005 | Current admission or past admission: MRDx, pre admit, secondary, admitting, transfer diagnosis |
| Alcohol use | Alcohol-related disorders [660] (2018 version) | Current admission: pre-admit |
| Hypertension | CIR007, CIR008 | Current admission or past admission: MRDx, pre admit, secondary, admitting, transfer diagnosis |
| Heart failure | CIR019 | Only past admissions: MRDx, pre admit, secondary, admitting, transfer diagnosis |
| Liver disease | DIG018, DIG019 | Only past admissions: MRDx, pre admit, secondary, admitting, transfer diagnosis |
| Renal Failure | GEN002, GEN003, GEN010 | Only past admissions: MRDx, pre admit, secondary, admitting, transfer diagnosis |

\*CCSR = Clinical Classifications Software Refined codes

## Notes

### Competing Interest Statement

The authors have declared no competing interest.

### Funding Statement

This study did not receive any funding.

### Author Declarations

Research Ethics Board approval was obtained from all participating hospitals. REB approval for University Health Network (Toronto), Sunnybrook Health Sciences Centre (Toronto), and St. Michael's Hospital (Toronto) was obtained through the integrated Clinical Trials Ontario platform, with St. Michael's Hospital as the "Board of Record". REB approval was also obtained from Trillium Health Partners (Mississauga) and Mount Sinai Hospital (Toronto). Clinical Trials Ontario Study ID 1394, https://www.ctontario.ca/

## REFERENCES

1. Singh TD, Fugate JE, Rabinstein AA. Central pontine and extrapontine myelinolysis: a systematic review. Eur J Neurol. Dec 2014;21(12):1443–50. doi:10.1111/ene.12571

2. Verbalis JG, Goldsmith SR, Greenberg A, et al. Diagnosis, evaluation, and treatment of hyponatremia: expert panel recommendations. Am J Med. Oct 2013;126(10 Suppl 1):S1–42. doi:10.1016/j.amjmed.2013.07.006

3. MacMillan TE, Cavalcanti RB. Outcomes in Severe Hyponatremia Treated With and Without Desmopressin. Am J Med. 03 2018;131(3):317.e1-317.e10. doi:10.1016/j.amjmed.2017.09.048

4. George JC, Zafar W, Bucaloiu ID, Chang AR. Risk Factors and Outcomes of Rapid Correction of Severe Hyponatremia. Clin J Am Soc Nephrol. 07 06 2018;13(7):984–992. doi:10.2215/CJN.13061117

5. Sterns RH. Treatment of Severe Hyponatremia. Clin J Am Soc Nephrol. 04 06 2018;13(4):641–649. doi:10.2215/CJN.10440917

6. MacMillan TE, Tang T, Cavalcanti RB. Desmopressin to Prevent Rapid Sodium Correction in Severe Hyponatremia: A Systematic Review. Am J Med. Dec 2015;128(12):1362.e15-24. doi:10.1016/j.amjmed.2015.04.040

7. Koul PA, Khan UH, Jan RA, et al. Osmotic demyelination syndrome following slow correction of hyponatremia: Possible role of hypokalemia. Indian J Crit Care Med. Jul 2013;17(4):231–3. doi:10.4103/0972-5229.118433

8. Verma AA, Guo Y, Kwan JL, et al. Patient characteristics, resource use and outcomes associated with general internal medicine hospital care: the General Medicine Inpatient Initiative (GEMINI) retrospective cohort study. CMAJ Open. Dec 11 2017;5(4):E842–E849. doi:10.9778/cmajo.20170097

9. Movig KL, Leufkens HG, Lenderink AW, Egberts AC. Validity of hospital discharge International Classification of Diseases (ICD) codes for identifying patients with hyponatremia. J Clin Epidemiol. Jun 2003;56(6):530–5. doi:10.1016/s0895-4356(03)00006-4

10. Woodfine JD, Sood MM, MacMillan TE, Cavalcanti RB, van Walraven C. Derivation and Validation of a Novel Risk Score to Predict Overcorrection of Severe Hyponatremia: The Severe Hyponatremia Overcorrection Risk (SHOR) Score. Clin J Am Soc Nephrol. 07 05 2019;14(7):975–982. doi:10.2215/CJN.12251018

11. Report prepared for the Office of the Information and Privacy Commissioner of Ontario in respect of PHIPA requirements for review and approval of prescribed persons and prescribed entities Institute for Clinical Evaluative Sciences (ICES); 2011.

